# An eight-state union history model with a transient phase after sexual debut and marriage

**DOI:** 10.64898/2026.10.28.26366604

**Authors:** Van Kinh Nguyen

## Abstract

Age of entry into the population at risk and the following union sequences are essential inputs for demographic and sexually transmitted infection models. These inputs are usually estimated separately from retrospective reports and current-status marital state. We fit the full union history as one joint-model of eight-state competing continuous time Markov chain using Demographic and Health Survey data from four countries with one to six survey rounds each. The fitted intensities are therefore estimated against the same exposure, structually producing consistent estimates for the proportion in every state. To account for the high first-year marriage rate after sexual debut, the coincident reports of debut and marriage age, and the never-marrying group, we reparameterise flexible hazard functions as log hazards, add a transient phase after debut and after marriage. Our model fitted well to both age and duration domain in all four country. In the first years after debut, the rate of marriage is from 1.7 (95% CI 1.0–2.9) times the later rate in Sierra Leone to 5.1 (95% CI 4.7–5.5) times in Rwanda. The raised rate lasts on average from 1.2 (95% CI 0.1–15.2) years in Sierra Leone to 3.5 (95% CI 2.3–5.2) years in Malawi. By 27 years after marriage, the widowed proportion reaches 23.3% (95% CI 22.2–25.1) in Rwanda, compared to 0.7% (95% CI 0.2–6.5) at 29 years in São Tomé, where the later-union proportion is 52.3% (95% CI 43.9–66.4). The divorced proportion stays flat over duration since marriage, 12.9% (95% CI 12.3–13.5) at 19 years in Rwanda. The premarital interval is thus a mixture of a short and a long component which should be taken into account in demographic and transmission model. As early marriage is followed by faster replacement of the first union, a cross-sectional proportion divorced would understate the amount of dissolution. Our model provide a framework for estimating a complete set of union-history rates to be used in transmission or a fertility models.

## Introduction

The age at first sex and the sequence of unions that follows set the exposure to a sexually transmitted infection and the time spent at risk of childbearing, so the same two quantities are estimated in epidemiology and in demography.^1,2^ Sexually transmitted infection models use the age of entry into the susceptible pool, the time spent in a stable union, and the states that follow the end of that union. These enter as rates of partnership formation and separation, in pair-formation form or as an individual history of unions,^1,3^ and mostly from survey means or from assumptions taken over from earlier work.

These behavioural inputs have been shown to be more important determinants of epidemic outcomes than disease biology.^4,5^ In urban Zambia and Rwanda, 55%–93% of new heterosexually acquired HIV infections among adults were estimated to occur inside serodiscordant marital or cohabiting partnerships.^6^ The states after a first union matter as much as the first union, since re-entry rate to the partnership pool after widowhood or after divorce is differ from first entry rate at the same age.^7,8^

Age at first marriage has risen across sub-Saharan Africa.^2,9,10^ The interval between sexual debut and first marriage has widened, and premarital fertility inside that interval has grown.^11,12^ Dissolution of a first union is frequent, with repartnering fast enough to keep the number separated at any one time small.^13–16^ Each quantity is however estimated on its own time scale: sexual debut rate fitted to age at first sex,^17^ age pattern of first marriage fitted to age at first marriage,^18,19^ a hazard of dissolution to the duration of the first union, and a hazard of repartnering to the time since that union ended. As a results, separately estimated quantities have to be assembled into a full chain of events, e.g., for a transmission model, and the assembled sequence is not guaranteed to reproduce the reported proportions in each state.

Multistate event-history methods with simultaneous-equation models of life events are available,^20,21^ but applications to marital histories are scarce. In the DHS, age at first sex and age at first marriage are reported retrospectively, each interval censored at the reported year of age, while marital state at interview is a current-status observation with no date attached. A survival model for one endpoint takes a event-time, so the dated reports and the undated state enter through different model likelihoods, requriring a joint modelling framework.^20^ In DHS data, a large part of the sample reports the same age for first sex and for first marriage, giving no debut date to define a hazard in duration since debut. Agreement between repeated reports of age at first sex is also poor.^17,22–24^

We fit the full union sequence as one eight-state continuous-time Markov chain per country, estimating sexual debut, first marriage, dissolution by separation and by widowhood, and entry into a later union together on survey data for Rwanda, Malawi, Sierra Leone and São Tomé and Príncipe. The four countries’ surveys span the information a country can offer this model, from six surveys with overlapping birth cohorts in Rwanda to a single survey in São Tomé; they also differ in recent history, with a sharply dated conflict in Sierra Leone, a mortality crisis in the mid-1990s in Rwanda and neither in Malawi.

## Methods

### Data

We used women’s recode files from nationally representative DHS surveys in Rwanda (six surveys), Malawi (four), Sierra Leone (three) and São Tomé and Príncipe (one). Each respondent reports an age at first sex and an age at first marriage, and the marital state at interview. A reported age *a* is taken as an event in [*a, a* + 1). Records are aggregated into weighted cells. A group is one combination of year of first marriage, age at first marriage, birth cohort and the longest duration observed in the survey. A row is one completed duration since marriage within a group, paired with the state observed at that duration, and a row’s weight is the summed survey weight of the respondents counted there. The group and row counts of each country are in eTable 1 in eAppendix A. Survey weights are propagated to the cell level, and intervals on observed proportions use the effective sample size of the DHS design, 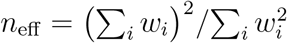 with *w*_*i*_ the weight of respondent *i* in the cell.^25,26^

### States, transitions and risk sets

The eight states are *V* never had sex, *X* sexually active and never married, *M* in a first marriage, *D*_1_ divorced from a first marriage, *W*_1_ widowed from a first marriage, *R* in a later union, and *D*_2_ and *W*_2_ the two ways a later union ends. The permitted transitions are *V ⟶ X, V ⟶ M* and *X ⟶ M* for the pre-marriage block, and *M* ⟶ *D*_1_, *M* ⟶ *W*_1_, *D*_1_ ⟶ *R, W*_1_ ⟶ *R, R* ⟶ *D*_2_, *R* ⟶ *W*_2_, *D*_2_ ⟶ *R* and *W*_2_ ⟶ *R* for the post-marriage block (Figure 1). Union order is in the state space because a later union dissolves at a different rate from a first one and is itself an outcome of the chain. A respondent in *V* is at risk of debut and of marriage at the same time, a respondent in *X* of marriage only.

**Figure 1:**
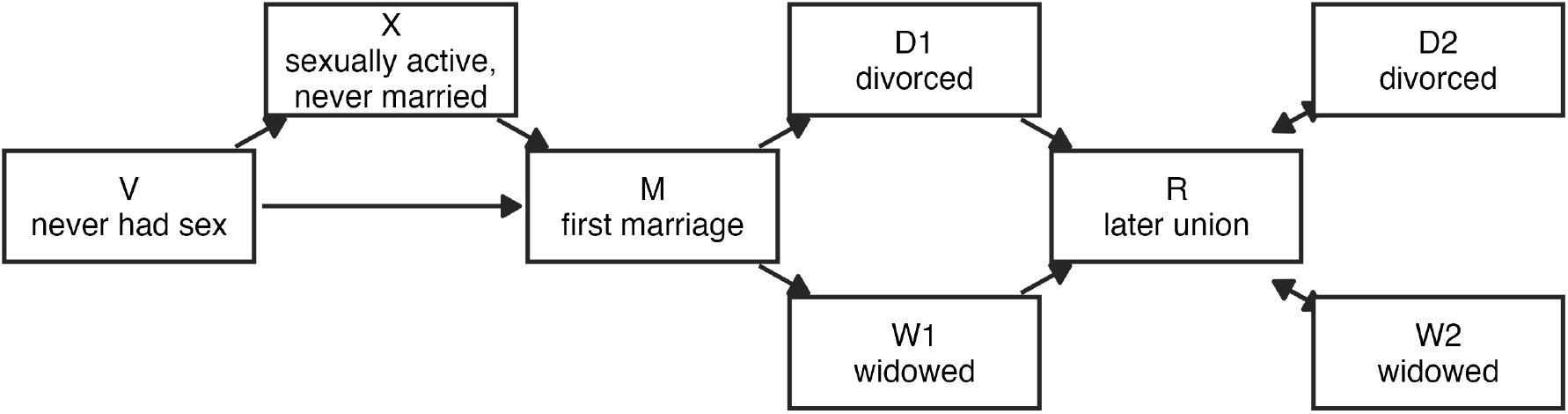
The eight states and the permitted transitions. The three transitions on the left form the pre-marriage block and the eight on the right the post-marriage block. Divorce and widowhood from a later union return to a later union, so the right-hand part of the chain is not absorbing.

### Duration since debut and the coincident reports

Two features of union formation are hard to represent in a chain whose intensities depend only on age. First, of the marriages reported after a debut in an earlier year, 39% (95% confidence interval 37–40) in Rwanda, 36% (95% CI 35–37) in Malawi, 24% (95% CI 23–25) in Sierra Leone and 50% (95% CI 45–54) in Sao Tomé fall in the first year after the debut, and the remainder are spread over the following decades. A hazard in duration since debut is defined only with a debut date at every age at risk, which is not available inside an episode that begins in *V*. The only value available is the age reported for a later year, applied to the whole episode, so the rate at duration zero is applied before any debut. Second, of those reporting both ages, first sex is reported in the year of marriage or in a later year by 84% (95% CI 83–85) in Rwanda, 65% (95% CI 64–66) in Malawi, 57% (95% CI 56–58) in Sierra Leone and 51% (95% CI 47–54) in São Tomé, so no time is observed between debut and marriage in those reports. Such reports enter the data with no exposure in *X*, so the only informative transition is *V ⟶ M*, and any term routing them through *X* competes with the debut curve for the same events. The numbers are weighted proportions with Jeffreys intervals on the effective sample size.

### A transient phase after debut

We applied the phase-type construction^27^ and split the sexually active unmarried state in two. A debut is entry to a transient phase *X*_1_, which is left for *X*_2_ at a constant rate ω, with marriage at risk in both phases,

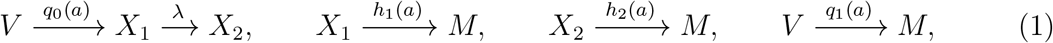

with 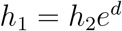 and one parameter *d* for how much more likely marriage is in the first phase. Entry to *X*_1_ is at the respondent’s own debut, so the duration dependence is defined without a debut date or a reported age. Among those still unmarried, the marriage intensity declines from *h*_1_ towards *h*_2_ with a time constant 1*/*ω. The chain is still Markov in age, so the observation model is unchanged.

The sinh-arcsinh shape on the log hazard^28^ is used for each of the four age-indexed intensities, with age *a*, peak location *µ*, width *σ* and asymmetry δ,

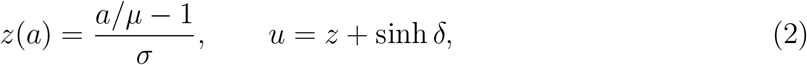

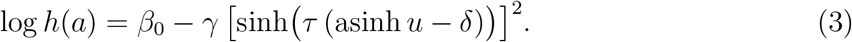

The four age-indexed intensities differ in where they peak and in how fast they fall away, so the location of the peak and the shape of the decline have to be set separately. The sinh-arcsinh form does that with an asymmetry parameter and a tail weight on top of a location-scale form,^28^ where in a form of fixed shape, such as the curve fitted to age at first marriage alone,^18^ the decline follows from the parameters that place the peak. At ages 45 to 49, 3.4% (95% CI 2.2–5.0) of the women in Rwanda, 0.3% (95% CI 0.1–0.9) in Malawi, 2.5% (95% CI 1.4–4.1) in Sierra Leone and 1.4% (95% CI 0.2–5.6) in São Tomé report no marriage, so a group that never marries has to be available under the model. A hazard built as a density over its own survivor function has a cumulative intensity that must diverge, which rules out a declining tail and leaves no group that never marries, while a log hazard falling as a power of age has a convergent cumulative intensity and keeps the never-marrying group of cure models.^29^ The shape is therefore imposed on the log hazard, where a steep decline of the hazard is permitted. The log hazard also fits the same events better in three of the four countries (eAppendix C).

In Equation 3, the sharpness *γ* and the tail exponent τ both set how fast the hazard falls away from its peak, once near the peak and once far from the peak. The curvature at the peak is proportional to *γτ*^2^, and far from the peak the log hazard falls as log *h* ∼ − (*γ*/4) exp(− 2 *τδ*)(2/*σ*)^2*τ*^ (*a/µ*)^2 *τ*^, a power of age with exponent 2 τ (eAppendix B). A curve observed over a limited range of age constrains those two combinations and not *γ* and τ apart. The curvature of the log hazard at its peak is therefore estimated in place of γ,

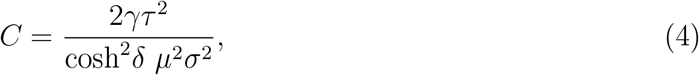

and *γ* follows from *C*. The curve near the peak then identifies *C* and the far tail identifies τ. Birth cohort is included as a linear shift of *µ* per decade. State *M* is absorbing within this block.

#### Post-marriage intensities

The post-marriage block is built on three intensities, divorce, widowhood and remarriage, and the later-union intensities are multiples of them, 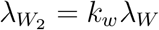 and 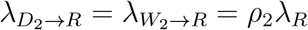. For a first marriage at age *a*_0_ in calendar year *t*_0_ observed *j* completed years later, with age *a* = *a*_0_ + *j* and calendar year *t* = *t*_0_ + *j*,

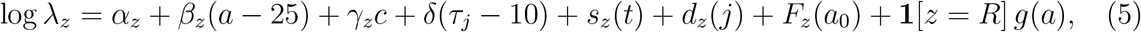

with *τ*_*j*_ = log(1 + *e*^*j*^) a smooth version of duration, finite in the first year, and *c* the birth year in decades from 1975. The slope *δ* is shared by the three intensities, while the intercept, the age slope and the cohort slope are free. The calendar term *s*_*z*_ and the duration term *d*_*z*_ are the two margins of a tensor array of penalised B-splines^30^ whose interaction block is set to zero, so the surface is additive. For parameter idenfiability, each spline basis enters with its intercept and linear part removed, and the constant and the linear-duration cells of the array are held at zero, so the linear plane in age, cohort and duration is represented by *β*_*z*_, *γ*_*z*_ and *δ* alone. *F*_*z*_ is a penalised smooth in age at first marriage and *g* a spline in current age, present for remarriage only given the proportion in a later union falls with age at first marriage, while the divorced and the widowed proportions change little (eAppendix D).

#### Likelihood and estimation

Intensities are assumed constant within a year of age or duration, so the transition matrix over one year is a matrix exponential of the generator, and the occupancy vector is propagated one year at a time from the entry state. The pre-marriage likelihood is taken per respondent. The four possible paths are no debut and no marriage, a debut and no marriage, a marriage in the year of the debut, and a debut followed by a marriage in a later year. In the order *V, X*_1_, *X*_2_, *M* the pre-marriage generator and the matrix over one year of age are

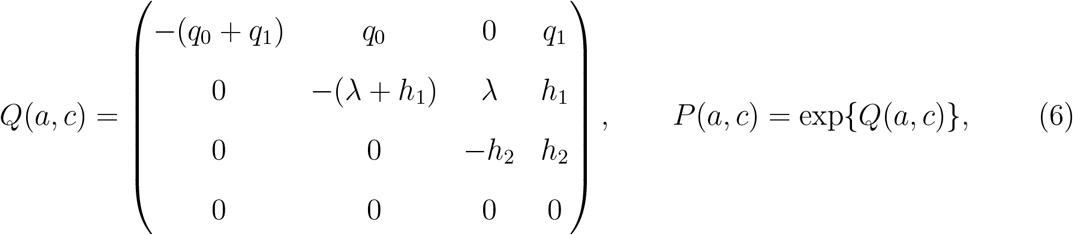

with *q*_0_, *q*_1_, *h*_1_ and *h*_2_ of Equation 1 evaluated at age *a* and cohort *c*. Writing *π*_*a*_ for the row vector of occupancy probabilities at age *a* and *m*_*a*_ for the vector that is one on the states the reported ages allow at the end of that year and zero elsewhere, the propagation is *π*_*a*+1_ = (π_*a*_*P*(*a, c*) ) ⊙ *m*_*a*_, *π*_1_ = *e*_*V*_, with ⊙ the elementwise product and *e*_*V*_ the unit vector on *V* . The contribution of respondent group *i* of weight *n*_*i*_ is *n*_*i*_ 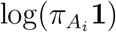, with *A*_*i*_ the age at which the reports end, i.e., age at first marriage or age at interview otherwise. In the order *M, D*_1_, *W*_1_, *R, D*_2_, *W*_2_ the post-marriage generator is

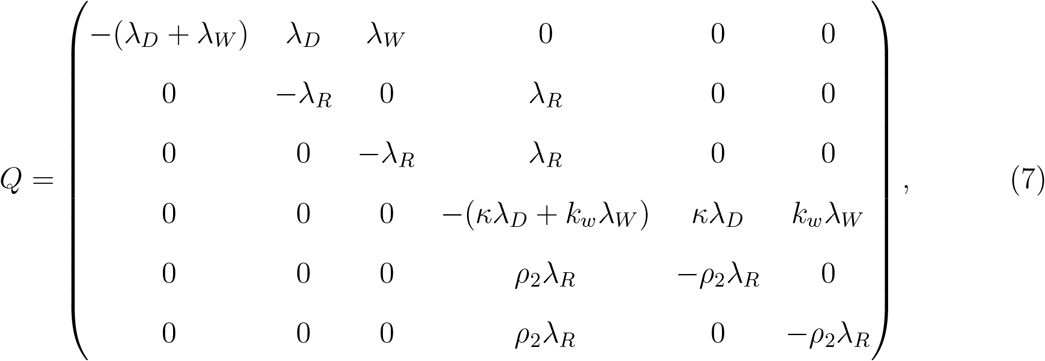

with the three intensities of Equation 5 evaluated at the age and the duration of the year in question. For cell *g* of weight *n*_*g*_, observed in state *Z*_*g*_ after *J*_*g*_ completed years since first marriage, the contribution is

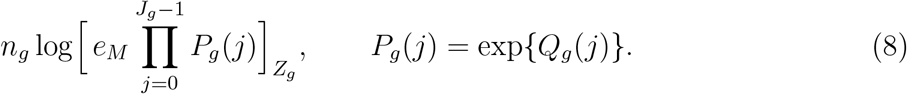

Each post-marriage observation is entered as the log probability of the state occupied at the interview duration. Confident intervals on the fitted quantities are calculated at the maximum likelihood in both blocks. All model comparisons use the log likelihood with the penalty contribution removed.^31^

#### Scoring

Models are judged on the margins measured in the data. For the pre-marriage block these are the annual probability of each transition by single year of age, and the annual probability of marriage by completed years since debut, with Jeffreys intervals^32^ on the observed proportions. For the post-marriage block, it is the proportion in each state by year of first marriage. Alongside the unpenalised AIC we report the binomial deviance of the fitted one-year cell probabilities against the observed counts, summed on each axis over cells with at least a hundredth of the exposure at risk in their panel.

#### Implementation

Models are written as C++ templates and fitted by maximum penalised likelihood with automatic differentiation in TMB^33^ from R.^34^ Every model is fitted from its own starting values.

## Results

### Union formation before marriage

The fitted and observed annual probability of each pre-marriage transition by single year of age is in Figure 2, and the probability of marriage by completed years since debut in Figure 3. On the age axis, the three transitions are reproduced in all four countries. Sexual debut rises to a peak between ages 17 and 20 and falls away, marriage in the year of debut peaks between 19 and 24, and marriage after debut peaks in the late teens and declines slowly. On the duration axis, the steep fall over the first three years and the long flat stretch after that fall are both reproduced in Rwanda, Malawi and Sierra Leone. In São Tomé, where exposure beyond fifteen years is scarce, the fitted curve runs below the high observed points but inside their 95% Jeffreys intervals.

**Figure 2:**
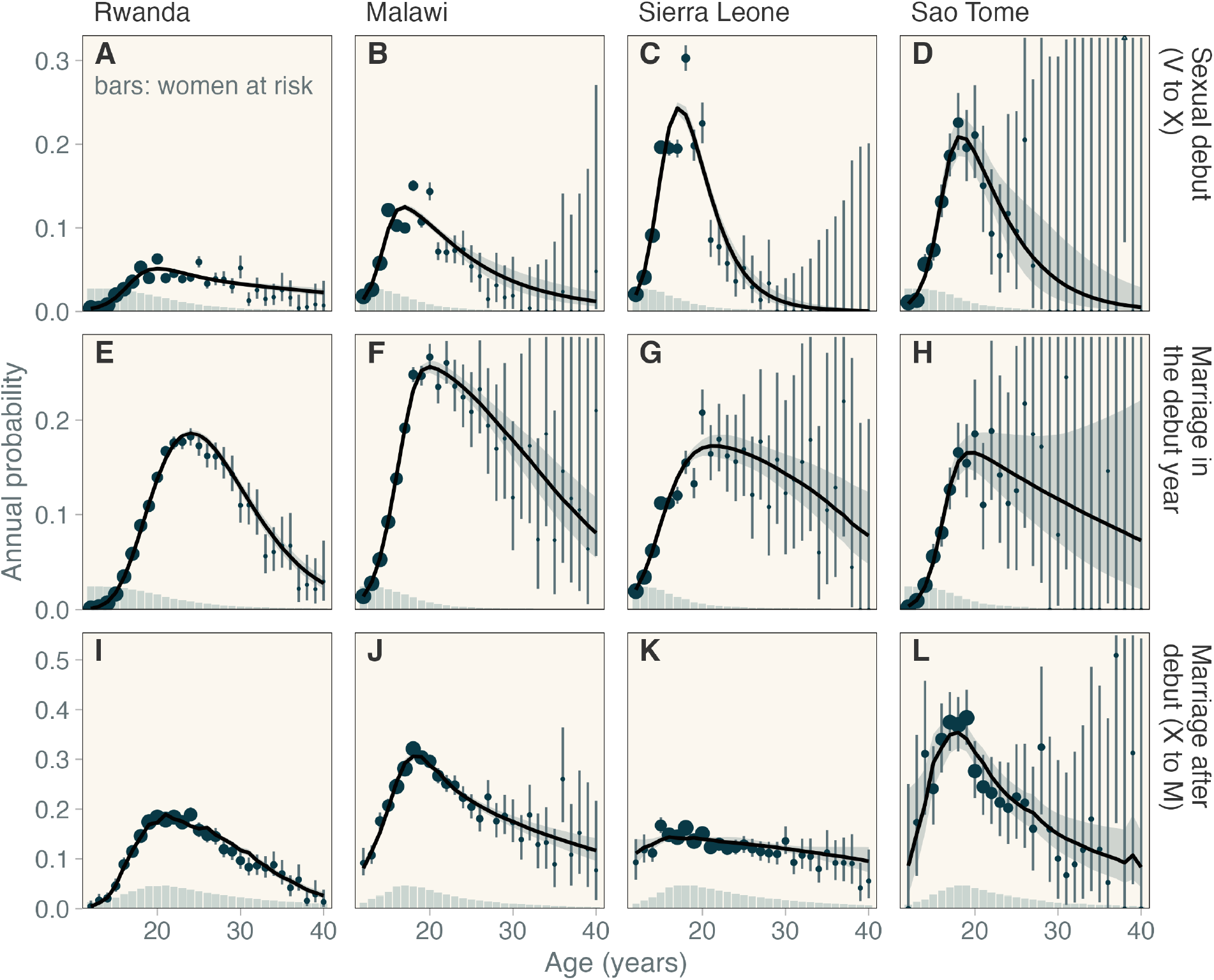
Pre-marriage transitions by single year of age, three transitions by row and four countries by column, under the adopted model. The line is the fitted annual probability and the shaded region the 95% CI of that probability. Observed points are drawn with a Jeffreys 95% interval and with area in proportion to the weighted number at risk, shown again by the bars on the panel floor on a square-root scale. An observed value above the panel limit is drawn as an open triangle on the edge.

**Figure 3:**
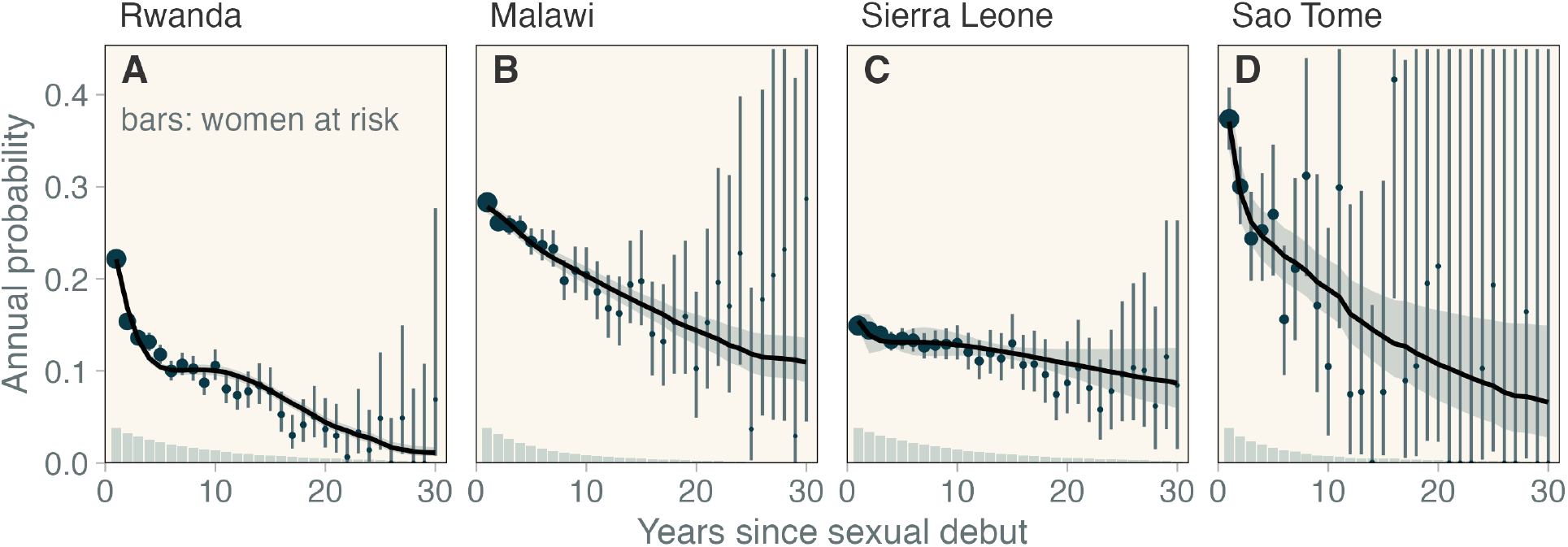
Marriage after sexual debut by completed years since debut, four countries, under the adopted model, with the 95% CI of the fitted curve.

### Union dissolution and remarriage

The observed and fitted proportion divorced, widowed or in a later union by years since first marriage is in Figure 4. The margin is reproduced in all four countries by the additive surface of Equation 5, including São Tomé with a single survey. The fitted interval overlaps the Jeffreys interval of the observed proportion in all 177 duration cells with at least 2% of their country’s weight. The three outcomes differ over duration since marriage. The widowed proportion accumulates steeply, from 0.4% (95% CI 0.3–0.6) in the first year after marriage to 23.3% (95% CI 22.2–25.1) at 27 years in Rwanda, 12.2% (95% CI 10.9–14.1) at 29 years in Malawi and 11.3% (95% CI 9.3–14.2) at 29 years in Sierra Leone. The divorced proportion is almost flat over the same span, 9.6% (95% CI 8.8–10.6) in the first year against 15.0% (95% CI 13.9–15.9) at 27 years in Rwanda, and 10.7% (95% CI 9.8–11.7) against 11.7% (95% CI 10.5–13.3) at 29 years in Malawi, since the outflow by remarriage is about as fast as the inflow by dissolution. The outflow appears in the later-union proportion, which rises to 12.8% (95% CI 12.2–14.2) at 27 years in Rwanda, 29.6% (95% CI 28.9–34.1) at 29 years in Malawi, 34.1% (95% CI 33.9–50.6) in Sierra Leone and 52.3% (95% CI 43.9–66.4) in São Tomé. Replacement of a first union is fastest in São Tomé and slowest in Rwanda. The widowed proportion in São Tomé is 0.7% (95% CI 0.2–6.5) at 29 years, the lowest of the four countries. On the margin of age at first marriage, the later-union proportion in Rwanda falls from 17.7% (95% CI 16.7–19.2) at age 16 to 5.3% (95% CI 4.8–6.0) at age 27, while the divorced and the widowed proportions change little over the same ages. Across marriage cohorts the widowed proportion falls from 23.2% (95% CI 21.6–25.8) in the 1975 cohort of Rwanda to 0.4% (95% CI 0.2–0.6) in the 2015 cohort, which is exposure, since few years are available after a recent marriage in which to be widowed (eAppendix D).

**Figure 4:**
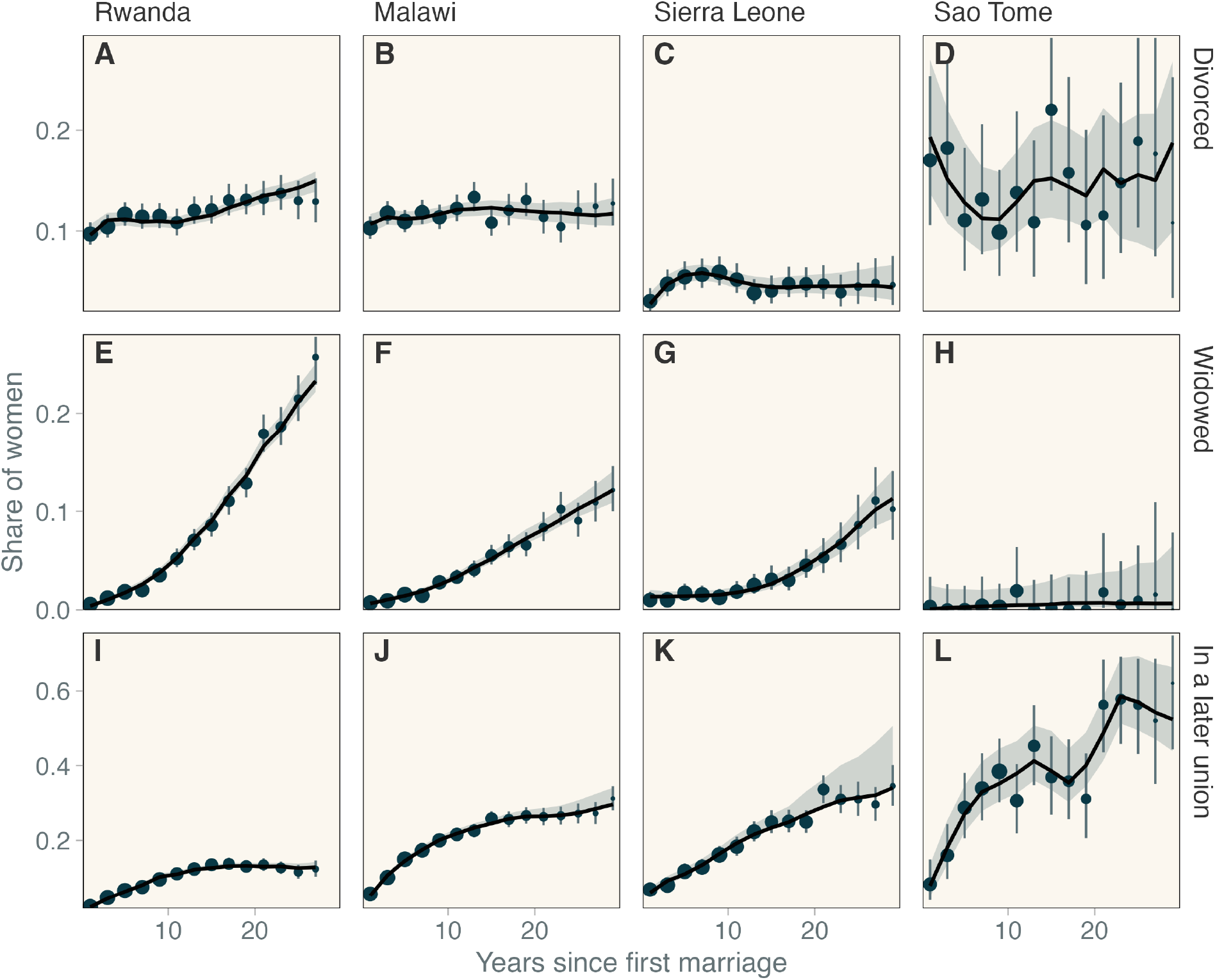
Observed and fitted proportion divorced, widowed or in a later union by years since first marriage, three outcomes by row and four countries by column, under the adopted model. The line is the fitted proportion and the shaded region the 95% CI of that proportion. Observed points are drawn with a Jeffreys 95% interval and with area in proportion to the weighted number observed. Two-year duration cells under 2% of a country’s weight are omitted.

### Model comparison

The model versions of both transition blocks are in Table 2. For the pre-marriage block, only the state space differs between the two models, since the single state after debut is the special case of Equation 1 at *d* = 0. The added phase reduces the unpenalised AIC in the four countries for one added parameter, and most of the gain is on the duration axis against a small loss on the age axis. For the post-marriage block, the additive surface improves on the parametric form with no surface in all four countries and comes close to a multinomial model of the proportions themselves (eAppendix E). An interaction between calendar time and duration improves the fit in Rwanda alone, where the survey rounds are most numerous. In São Tomé, with one survey round, the more complex models fit worse or do not converge.

**Table 1:** Parameters of the adopted model with a direct interpretation, estimate and 95% confidence interval. The peak age is in years, for the 1975 birth cohort, and the curvature at the peak is per year squared. The multiplier and the mean time belong to the first phase after debut. The duration slope is per year and is shared by the three intensities after marriage, and the last three rows are the multipliers of a later union. Every parameter of both blocks is in eAppendix F.

|  | Rwanda | Malawi | S. Leone | São Tomé |
| --- | --- | --- | --- | --- |
| *Sexual debut* |  |  |  |  |
| Peak age | 21.3<br>(21.0–21.7) | 17.6<br>(17.3–17.9) | 17.4<br>(17.2–17.6) | 18.4<br>(17.4–19.4) |
| Curvature | 0.032<br>(0.027–0.039) | 0.073<br>(0.062–0.086) | 0.147<br>(0.134–0.162) | 0.139<br>(0.060–0.318) |
| *Marriage in the year of debut* |  |  |  |  |
| Peak age | 23.7<br>(23.5–23.9) | 19.8<br>(18.9–20.7) | 21.2<br>(19.4–23.1) | 25.2<br>(19.2–33.0) |
| Curvature | 0.030<br>(0.028–0.032) | 0.021<br>(0.014–0.030) | 0.008<br>(0.003–0.018) | 0.009<br>(0.001–0.070) |
| *Marriage after debut* |  |  |  |  |
| Peak age | 24.7<br>(24.1–25.4) | 18.8<br>(18.1–19.5) | 15.7<br>(13.9–17.7) | 18.0<br>(16.3–20.0) |
| Curvature | 0.016<br>(0.013–0.020) | 0.031<br>(0.017–0.059) | 0.004<br>(0.001–0.013) | 0.035<br>(0.009–0.135) |
| *The transient phase* |  |  |  |  |
| Marriage multiplier | 5.1<br>(4.7–5.5) | 2.0<br>(1.8–2.1) | 1.7<br>(1.0–2.9) | 2.6<br>(1.7–3.9) |
| Mean time (years) | 2.5<br>(2.1–2.9) | 3.5<br>(2.3–5.2) | 1.2<br>(0.1–15.2) | 1.3<br>(0.5–2.9) |
| *After marriage* |  |  |  |  |
| Duration slope | –0.073<br>(–0.108 to –0.038) | –0.090<br>(–0.120 to –0.059) | –0.123<br>(–0.179 to –0.068) | –0.069<br>(–0.159 to 0.021) |
| Dissolution $\kappa$ | 6.3<br>(4.4–8.9) | 3.7<br>(2.7–5.2) | 1.3<br>(0.59–2.8) | 46<br>(14–150) |
| Widowhood $k_w$ | 1.3<br>(1.1–1.5) | 1.2<br>(0.95–1.6) | 0.69<br>(0.46–1.0) | 0.67<br>(0.08–5.8) |
| Repartnering $\rho_2$ | 1.1<br>(0.51–2.2) | 1.6<br>(0.97–2.5) | 0.92<br>(0.34–2.5) | 15<br>(6.3–38) |

**Table 2:** Unpenalised AIC of the model versions of the two transition blocks, by country, with the number of parameters per country. Pre-marriage rows are scored on the per-respondent likelihood of this paper, and the deviance rows below them give the contribution of each time axis to that likelihood. Lower is better in every column.

| Specification | Parameters<sup>a</sup> | Rwanda | Malawi | S. Leone | São Tomé |
| --- | --- | --- | --- | --- | --- |
| *Before marriage* |  |  |  |  |  |
| One state after debut | 22 | 295,224.6 | 278,119.1 | 127,482.3 | 11,660.6 |
| Transient phase after debut | 23 | 294,000.3 | 277,882.7 | 127,453.7 | 11,641.4 |
| *Before marriage, deviance by axis* |  |  |  |  |  |
| Duration, one state |  | 915.6 | 163.7 | 30.5 | 41.6 |
| Duration, with the phase |  | 66.7 | 18.2 | 19.3 | 23.8 |
| Age, change with the phase |  | -20.8 | +87.6 | -4.7 | +0.4 |
| *After marriage* |  |  |  |  |  |
| Parametric, no surface | 16 | 57,229.4 | 63,545.3 | 19,087.3 | 2,555.5 |
| Additive surface | 136 | 56,152.6 | 62,967.8 | 18,872.2 | 2,533.2 |
| Blocked tensor interaction | 262 | 56,068.8 | 62,967.4 | 18,872.2 | not fitted<sup>b</sup> |
| Free tensor interaction | 892 | 56,062.5 | 62,982.6 | 18,884.0 | 2,536.9 |
| Multinomial model<sup>c</sup> |  | 56,031.9 | 62,905.8 | 18,846.8 | 2,567.5 |

<sup>a</sup> Post-marriage counts are those of Rwanda, Malawi and Sierra Leone, with three fewer in every post-marriage row of São Tomé, where the cohort term is dropped.
<sup>b</sup> The blocked interaction did not converge in São Tomé from any of six starting values, among them the additive fit, the free-interaction fit and the additive fit under wider bounds.
<sup>c</sup> A multinomial logistic regression of the six states, written as five nested binary generalised additive models and fitted to the same cells and weights with no chain and no intensities (eAppendix E). The model is penalised, so its parameter count is an effective degrees of freedom that differs by country.

## Discussion

We fitted the union history from sexual debut to the end of a later union as one eight-state chain per country. The estimated marriage rate after debut depends on the time since the debut. The age pattern, the steep fall of the marriage rate over the first years and the long flat stretch at later durations are reproduced by one transient state in the chain. In the first phase, marriage is 5.1 (95% CI 4.7–5.5) times more likely than later in Rwanda. The ratio of the two rates is above one in all four countries. The transient phase from the debut to the lower marriage rate lasts on average 3.5 years (95% CI 2.3–5.2) in Malawi and between one and two and a half years elsewhere. The premarital interval is therefore a mixture of a short and a long component in all four countries. For a transmission model, the exposure of the unmarried follows from the mixture and not from its mean.

The transient phase can be interpreted as a pending union formalisation. Where marriage after debut formalises a union begun at or near the debut, the first phase covers the period before the formalisation. The rate of leaving the first phase is then the rate at which an unformalised union becomes an ordinary premarital interval. The reported coincidence of the two ages is the same mechanism at duration zero. Those reports stay on the direct transition from never had sex to married.@ai add relevant statistics and citations@ Two consequences follow for the shape of the hazard. First, the sharpness of a curve near its peak and the power of age far from the peak are separate quantities. The fitted curvature at the peak differs by more than an order of magnitude between transitions within one country, with no overlap of the two intervals (Table 1). One value of either quantity across the three transitions is therefore not available. The curvature at the peak is estimated directly here, and the sharpness is derived from the curvature through Equation 4. Second, a never-marrying group is possible only under a declining tail. A declining tail follows from a shape imposed on the log hazard and not from a density. A never-marrying group is needed in all four countries.

The four countries differ in their fitted patterns. Sexual debut peaks three to four years later in Rwanda than in Sierra Leone. The shift towards an earlier debut across birth cohorts is material only in Rwanda (eTable 3 in eAppendix F). After marriage, the contrast is between widowhood and replacement of the union. Widowhood reaches 23.3% by 27 years of marriage in Rwanda and stays below 1% in São Tomé, where instead half of first unions have been replaced by 29 years (Figure 4). The divorced proportion is almost flat in Rwanda and in Malawi (Figure 4), since the outflow by remarriage is about as fast as the inflow by dissolution. A cross-sectional proportion divorced therefore understates the amount of dissolution. Early marriage is followed by faster replacement of the first union, with no more dissolution of the first union (eAppendix D).

The pattern agrees with what has been reported for the region. Rapid remarriage after divorce is documented in rural Malawi^13^ and in Ethiopia.^14^ Dissolution and repartnering have been described across sub-Saharan Africa from Demographic and Health Survey data,^15,16^ as have the regional trends in age at marriage and at first birth.^2,10,12^ The contribution here is a joint fit of the three outcomes as competing destinations of one chain. The flat divorced proportion and the rising later-union proportion then follow from one set of intensities.

For a transmission model, the added phase changes the exposure window as well as the fit. An age-indexed hazard gives the same marriage rate at one year since debut and at ten, so the premarital interval is close to exponential. The interval mean is then the only quantity available to a pair-formation rate.^1^ Under the added phase, the same mean splits into a short interval before a pending formalisation and a long interval for the remainder. Most of the premarital exposure falls in the long interval. The same holds after a first union ends, where the rate of repartnering sets the time spent in the partnership pool. ^7,8^ Calibration to the mean of a mixed interval misplaces the exposure. Projections for the same setting from independent models still diverge,^5,35^ with inputs of partnership formation and separation among the traced sources of the disagreement.^4^

### Strengths and limitations

The whole union history is fitted as one state space with competing destinations. The outcomes after marriage are therefore estimated against the same exposure. The proportion in every state follows from the fitted intensities. The design uses public survey data and the same procedure in all four countries. Three limitations remain. (i) Reporting bias is documented for age at first sex^36^ and may be present in age at first marriage as well. Neither is estimated here. (ii) The post-marriage data are current status, so the intensities are identified from the composition of state occupancies and not from observed transitions. Remarriage after widowhood is not separated from remarriage after divorce, since the surveys do not record how the earlier union ended for a respondent now in a later union. (iii) The later-union multipliers κ, *k*_*w*_ and ρ_2_ scale the dissolution and the repartnering rates of a later union. Without them, a later union would dissolve at the rate of a first union, against the fitted state proportions. The multipliers are identified from the composition of state occupancies alone. The model holds no time scale for duration in a later union and no term for selection into repartnering, so each multiplier absorbs the two omissions. Each multiplier is therefore interpreted as the correction parameter and not as a ratio of risks.

Before marriage, the duration dependence is supplied by one transient state at a cost of one parameter per country, and the gain in goodness of fit is large in all four countries. After marriage, an additive calendar-by-duration surface with a penalised smooth in age at first marriage comes close to a multinomial model of the proportions themselves. The chain remains identified in a country with a single survey. The fitted intensities can enter a transmission or a fertility model directly, in place of a single mean duration.

## Data Availability

The data are the women's recode files of the
Demographic and Health Surveys. Any researcher can obtain them from the DHS Program
at \url{https://dhsprogram.com} by registering an account and submitting a request
that states the intended use, which the Program approves without cost; the survey
rounds used here are named in the Methods. No file was altered before use, and the
code named below reproduces the analysis cells from the downloaded recode files.

https://dhsprogram.com

## Notes

### Competing Interest Statement

The authors have declared no competing interest.

### Author Declarations

The study used ONLY openly available human data that were originally located at the DHS Program at https://dhsprogram.com.

