## Appendix for "An eight-state union history model with a transient phase after sexual debut and marriage"

### eAppendix A. Cell counts

The post-marriage likelihood runs over the groups and rows of eTable 1, defined in the Methods.

eTable 1: Surveys, groups, rows and weight of the post-marriage likelihood by country. The weight is the sum of the survey weights over the rows.

| Country | Surveys | Groups | Rows | Weight |
|---|---|---|---|---|
| Rwanda | 6 | 1,091 | 10,728 | 29,533 |
| Malawi | 4 | 1,105 | 10,954 | 30,178 |
| Sierra Leone | 3 | 990 | 6,714 | 11,587 |
| São Tomé | 1 | 398 | 938 | 1,182 |

### eAppendix B. Identifiability of the peak and the tail

In Equation 3 the sharpness $\gamma$ and the tail power $\tau$ act in tandem. A hazard curve over a limited observed range of age constrains the combinations, making $\gamma$ and $\tau$ weakly separated. Write the shape of Equation 3 as $\log h(a) = \beta_0 - \gamma\,[\sinh(\tau(\operatorname{asinh} u - \delta))]^2$ with $u = z + \sinh\delta$ and $z = (a/\mu - 1)/\sigma$. The squared term vanishes at $\operatorname{asinh} u = \delta$, at $u = \sinh\delta$, $z = 0$ and $a = \mu$. The peak hazard is therefore at $a = \mu$.

Close to the peak the log hazard is a parabola in age. Put $w = \operatorname{asinh} u - \delta$. For small $w$, $\sinh(\tau w) \approx \tau w$, and $\mathrm{d}w/\mathrm{d}a = 1/(\mu\sigma\sqrt{1+u^2}) = 1/(\mu\sigma\cosh\delta)$ at the peak. Hence $\log h(a) \approx \beta_0 - \gamma\tau^2(a-\mu)^2/(\mu^2\sigma^2\cosh^2\delta)$. The second derivative at the peak is $-C$ with $C$ the curvature of Equation 4.

Far from the peak the log hazard falls as a power of age. For large $u$, $\operatorname{asinh} u \approx \log(2u)$ and $\sinh(x) \approx e^x/2$, so the bracket tends to $\frac{1}{2}(2u)^\tau e^{-\tau\delta}$ and

$$\log h(a) - \beta_0 \;\longrightarrow\; -\frac{\gamma}{4}\, e^{-2\tau\delta}\left(\frac{2}{\sigma}\right)^{2\tau}\left(\frac{a}{\mu}\right)^{2\tau}. \tag{9}$$

The power of age is $2\tau$. The expression describes the shape of the decline.

The curvature at the peak and the coefficient of the tail are different mixtures of $\gamma$ and $\tau$. The curvature $C$ is estimated in place of $\gamma$, and $\gamma$ follows from $C$ through Equation 4. The shape near the peak is then set by $C$ and the decline in the tail by $\tau$.

### eAppendix C. The density hazard and the log hazard

The same four-state chain was fitted with the sinh-arcsinh shape on the density and with the shape on the log hazard, state $M$ absorbing, and scored on the episode likelihood of the single state after debut. The log hazard is ahead in Rwanda, Malawi and Sierra Leone and slightly behind in São Tomé, where the number of respondents is smallest.

eTable 2: Unpenalised AIC of the sinh-arcsinh density hazard and of the sinh-arcsinh log hazard, state $M$ absorbing, scored on the episode likelihood of the single state after debut.

| Country | Density hazard | Log hazard | Difference |
|---|---|---|---|
| Rwanda | 294,702.9 | 294,306.5 | −396.4 |
| Malawi | 278,221.3 | 278,018.4 | −203.0 |
| Sierra Leone | 127,680.9 | 127,474.0 | −206.9 |
| São Tomé | 11,644.7 | 11,646.8 | +2.1 |

### eAppendix D. The other margins after marriage

The fitted model is read on two further margins of the data. eFigure 1 is by year of first marriage. The widowed proportion falls from 23.2% (95% CI 21.6–25.8) in the 1975 marriage cohort of Rwanda to 0.4% (95% CI 0.2–0.6) in the 2015 cohort, from 12.3% (95% CI 10.8–15.1) in 1970 to 0.7% (95% CI 0.6–1.1) in 2010 in Malawi, and from 9.7% (95% CI 8.1–12.6) in 1980 to 1.1% (95% CI 0.7–1.9) in 2015 in Sierra Leone. The later-union proportion falls with the widowed one. Both declines are exposure, since few years are available after a marriage in 2010 in which to be widowed or to remarry.

eFigure 2 is by age at first marriage, so that marriages at child ages and marriages at adult ages with the same survey date are shown apart. Averaged over duration, the divorced proportion in Rwanda falls from 15.7% (95% CI 14.5–17.0) at age 16 at first marriage to 11.2% (95% CI 10.3–12.2) at age 27, and the widowed proportion from 12.0% (95% CI 11.1–13.1) to 6.3% (95% CI 5.8–7.2) over the same ages. The later-union proportion falls in Rwanda from 17.7% (95% CI 16.7–19.2) at age 16 to 5.3% (95% CI 4.8–6.0) at age 27, in Malawi from 29.9% (95% CI 29.1–32.5) at age 14 to 15.9% (95% CI 15.3–17.1) at age 23, and in Sierra Leone from 25.2% (95% CI 24.6–32.3) at age 14 to 16.5% (95% CI 16.2–19.5) at age 20. Early marriage is therefore followed by faster replacement of the first union, with no more dissolution of that union.

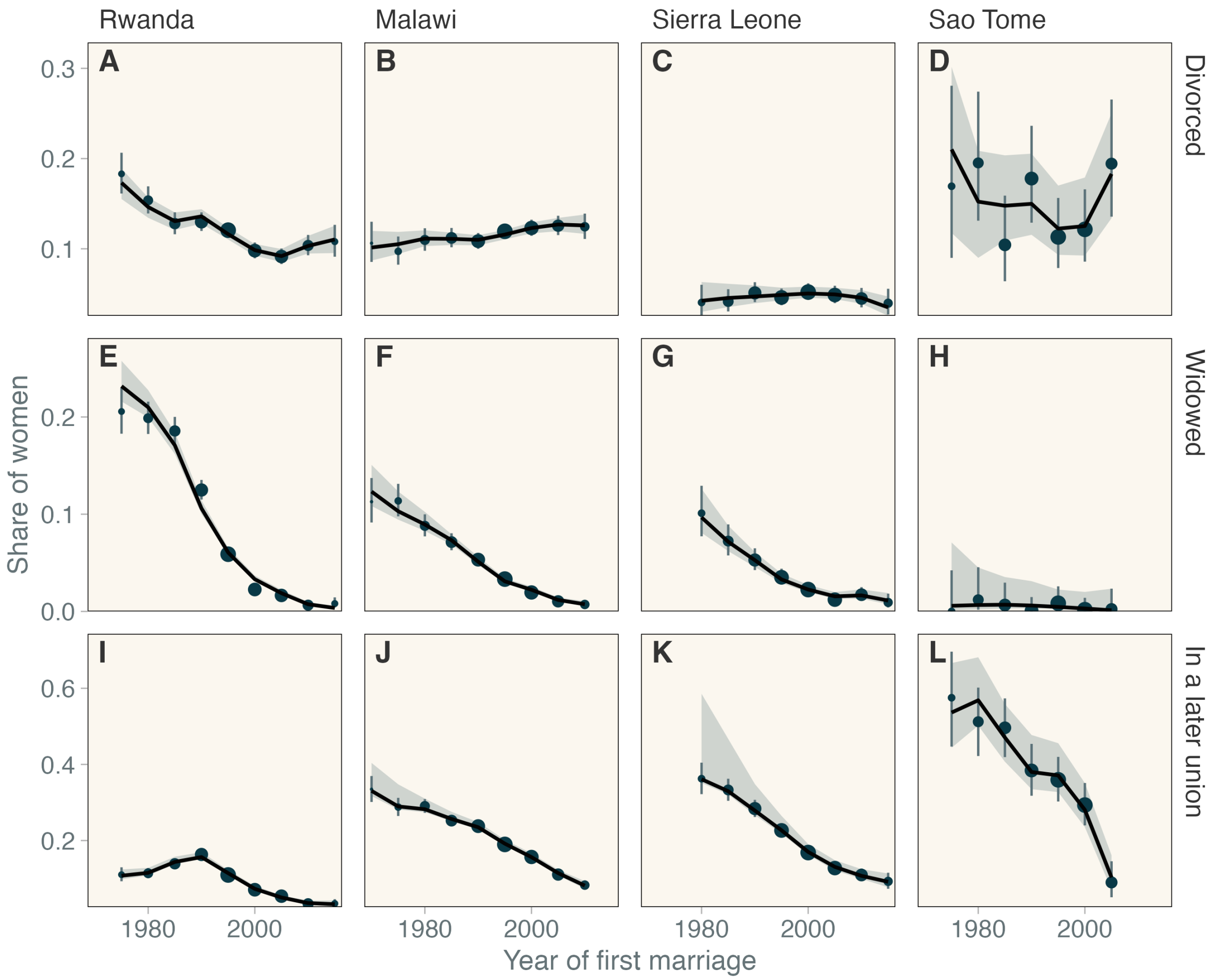


eFigure 1: Observed and fitted proportion divorced, widowed or in a later union by year of first marriage, three outcomes by row and four countries by column, under the adopted model. Devices are those of Figure 4 in the Results, with five-year marriage cohorts under 2% of a country's weight omitted.

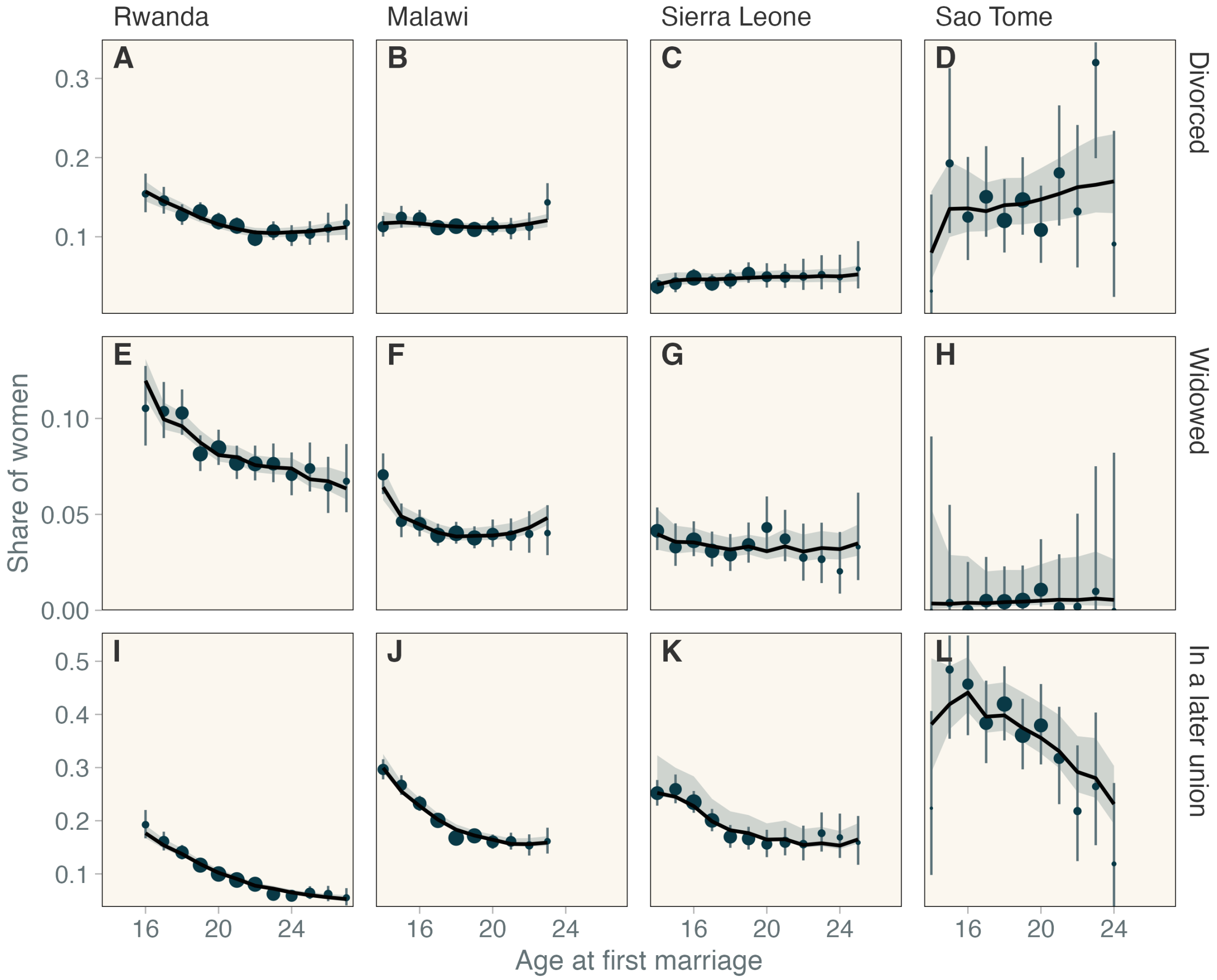


eFigure 2: Observed and fitted proportion divorced, widowed or in a later union by age at first marriage, three outcomes by row and four countries by column, under the adopted model. Devices are those of Figure 4 in the Results, with single years of age under 2% of a country's weight omitted.

### eAppendix E. The multinomial model of the proportions

The six states of the last row of Table 2 in the Results are written as five nested binary contrasts, i.e., a first union against any ended union, a later union against a first union that ended, widowhood against divorce in the first union, a later union in progress against a later union that ended, and widowhood against divorce in the later union. Each contrast is fitted as a binomial generalised additive model by restricted maximum likelihood in `mgcv`.[32] Two eTable 3: Pre-marriage parameters of the adopted phase model, estimate and 95% confidence interval. The level of each intensity is reported at its peak age, the peak age itself for the 1975 birth cohort, and the cohort term as the ratio of the peak age per decade of birth cohort.

| | Rwanda | Malawi | Sierra Leone | São Tomé |
|---|---|---|---|---|
| *Sexual debut,* $q_0$ | | | | |
| Peak age | 21.3 (21.0–21.7) | 17.6 (17.3–17.9) | 17.4 (17.2–17.6) | 18.4 (17.4–19.4) |
| Width $\sigma$ | 0.14 (0.12–0.16) | 0.14 (0.10–0.18) | 0.21 (0.17–0.26) | 0.11 (0.01–0.87) |
| Asymmetry $\delta$ | 1.27 (1.14–1.41) | 1.05 (0.91–1.20) | 0.67 (0.55–0.79) | 0.71 (0.35–1.06) |
| Curvature at the peak $C$ (per year$^2$) | 0.032 (0.027–0.039) | 0.073 (0.062–0.086) | 0.147 (0.134–0.162) | 0.139 (0.060–0.318) |
| Tail exponent $\tau$ | 0.12 (0.03–0.45) | 0.36 (0.26–0.48) | 0.16 (0.04–0.66) | 0.49 (0.12–1.89) |
| Peak age per decade of cohort (ratio) | 0.967 (0.964–0.971) | 0.999 (0.996–1.001) | 0.994 (0.990–0.997) | 0.996 (0.984–1.009) |
| Intensity at the peak (per year) | 0.072 (0.070–0.075) | 0.190 (0.183–0.198) | 0.345 (0.327–0.363) | 0.374 (0.324–0.432) |
| *Marriage without a reported debut,* $q_1$ | | | | |
| Peak age | 23.7 (23.5–23.9) | 19.8 (18.9–20.7) | 21.2 (19.4–23.1) | 25.2 (19.2–33.0) |
| Width $\sigma$ | 0.27 (0.23–0.31) | 0.05 (0.01–0.17) | 0.19 (0.10–0.36) | 0.20 (0.03–1.30) |
| Asymmetry $\delta$ | 1.27 (1.04–1.51) | 0.84 (0.72–0.97) | 0.96 (0.59–1.32) | 1.60 (−0.43 to 3.62) |
| Curvature at the peak $C$ (per year$^2$) | 0.030 (0.028–0.032) | 0.021 (0.014–0.030) | 0.008 (0.003–0.018) | 0.009 (0.001–0.070) |
| Tail exponent $\tau$ | 0.22 (0.02–2.80) | 0.89 (0.82–0.97) | 1.05 (0.73–1.51) | 0.63 (0.02–17.41) |
| Peak age per decade of cohort (ratio) | 1.056 (1.054–1.059) | 1.035 (1.032–1.038) | 1.068 (1.061–1.075) | 1.027 (0.996–1.059) |
| Intensity at the peak (per year) | 0.199 (0.194–0.204) | 0.282 (0.272–0.293) | 0.188 (0.171–0.205) | 0.131 (0.088–0.194) |
| *Marriage in the second phase,* $h_2$ | | | | |
| Peak age | 24.7 (24.1–25.4) | 18.8 (18.1–19.5) | 15.7 (13.9–17.7) | 18.0 (16.3–20.0) |
| Width $\sigma$ | 0.38 (0.15–0.97) | 0.06 (0.03–0.12) | 0.12 (0.04–0.34) | 0.13 (0.04–0.40) |
| Asymmetry $\delta$ | 1.54 (0.29–2.78) | 0.98 (0.78–1.18) | 2.42 (0.76–4.09) | 1.15 (0.28–2.02) |
| Curvature at the peak $C$ (per year$^2$) | 0.016 (0.013–0.020) | 0.031 (0.017–0.059) | 0.004 (0.001–0.013) | 0.035 (0.009–0.135) |
| Tail exponent $\tau$ | 1.32 (0.70–2.51) | 0.61 (0.45–0.84) | — | 0.24 (0.02–3.38) |
| Peak age per decade of cohort (ratio) | 1.095 (1.083–1.107) | 1.001 (0.993–1.009) | 1.398 (1.131–1.729) | 1.045 (0.984–1.110) |
| Intensity at the peak (per year) | 0.129 (0.120–0.138) | 0.264 (0.238–0.293) | 0.147 (0.124–0.174) | 0.319 (0.257–0.398) |
| *The transient phase* | | | | |
| Marriage multiplier in the first phase $e^d$ | 5.11 (4.71–5.55) | 1.95 (1.80–2.11) | 1.72 (1.02–2.91) | 2.58 (1.71–3.89) |
| Mean time in the first phase $1/\lambda$ (years) | 2.47 (2.10–2.90) | 3.47 (2.32–5.19) | 1.24 (0.10–15.16) | 1.25 (0.54–2.90) |

forms were fitted, the three-way tensor product of duration since first marriage, year of first marriage and age at first marriage that is in the table, and an additive form, a tensor product of duration and year plus a smooth in age at first marriage.

### eAppendix F. Parameter estimates

The pre-marriage parameters of the adopted phase model are in eTable 3, and the parametric terms after marriage in eTable 4. Each estimate is given with a 95% confidence interval Monotone transformed quantities, such as the peak age $e^{\mu}$ or the mean time in the first phase $1/\lambda$, has its interval transformed with it. One tail exponent of the second-phase marriage intensity in Sierra Leone has no standard error since the observed information is not positive definite in that coordinate. After marriage the three cohort slopes are fixed at zero in São Tomé, where one survey does not separate cohort from calendar time.

eTable 4: Parametric terms of the post-marriage intensities, estimate and 95% confidence interval. The intercept is the log intensity at age 25 and duration 10 with the penalised terms at zero, which is a reference point and not a fitted rate.

| | Rwanda | Malawi | Sierra Leone | São Tomé |
|---|---|---|---|---|
| *Intercept $\alpha_z$, log intensity per year at age 25 and duration 10* | | | | |
| Divorce | −5.49 (−5.84 to −5.14) | −4.57 (−4.74 to −4.40) | −4.99 (−5.49 to −4.49) | −5.08 (−6.39 to −3.76) |
| Widowhood | −8.06 (−8.47 to −7.66) | −4.50 (−4.70 to −4.31) | −4.90 (−5.27 to −4.52) | −7.38 (−9.55 to −5.21) |
| Later union | −9.40 (−9.72 to −9.07) | −0.38 (−0.57 to −0.19) | −2.73 (−3.12 to −2.34) | 6.45 (5.44–7.47) |
| *Age slope $\beta_z$, per year* | | | | |
| Divorce | 0.003 (−0.015 to 0.022) | 0.066 (0.045–0.086) | 0.055 (−0.005 to 0.115) | 0.036 (−0.012 to 0.083) |
| Widowhood | 0.403 (0.365–0.441) | 0.156 (0.126–0.187) | −0.004 (−0.034 to 0.026) | 0.072 (−0.094 to 0.239) |
| Later union | 1.644 (1.594–1.695) | −0.209 (−0.244 to −0.174) | 0.189 (0.117–0.262) | −2.642 (−2.774 to −2.511) |
| *Cohort slope $\gamma_z$, per decade* | | | | |
| Divorce | 0.165 (0.058–0.273) | 0.245 (0.078–0.412) | 0.332 (−0.177 to 0.841) | not fitted |
| Widowhood | 3.171 (2.913–3.430) | 0.755 (0.567–0.942) | −1.147 (−1.421 to −0.873) | not fitted |
| Later union | 0.512 (0.377–0.648) | 1.529 (1.359–1.700) | 1.105 (0.608–1.602) | not fitted |
| *Shared terms* | | | | |
| Duration slope $\delta$, per year | −0.073 (−0.108 to −0.038) | −0.090 (−0.120 to −0.059) | −0.123 (−0.179 to −0.068) | −0.069 (−0.159 to 0.021) |
| Dissolution, later union $\kappa$ | 6.28 (4.44–8.90) | 3.74 (2.67–5.24) | 1.28 (0.59–2.77) | 45.60 (13.84–150.23) |
| Widowhood, later union $k_w$ | 1.27 (1.10–1.47) | 1.24 (0.95–1.62) | 0.69 (0.46–1.03) | 0.67 (0.08–5.83) |
| Repartnering, later union $\rho_2$ | 1.05 (0.51–2.19) | 1.57 (0.97–2.53) | 0.92 (0.34–2.46) | 15.34 (6.26–37.60) |
